# Changes in dental care behaviour between 2002 and 2012 and its association with complete dentition in men and women in Switzerland

**DOI:** 10.1101/2019.12.13.19014878

**Authors:** Cornelia Schneider, Nicola U. Zitzmann, Elisabeth Zemp

## Abstract

**Background:** In industrialized countries, the awareness of oral hygiene measures has increased and the number of missing teeth has been decreasing. A higher number of missing teeth was reported by women despite their more intense oral hygiene. The aim of this study was to compare oral hygiene and its association to oral health with a complete dentition in women and men in Switzerland between 2002 and 2012.

**Methods:** Weighted data from the Swiss-Health-Surveys in 2002 and 2012 were used to quantify the number of missing teeth, the prevalence of prosthetic dental restorations, dental visits and tooth brushing. Sex-stratified logistic regression analysis was performed for subjects aged ≥65-yrs to assess associations between a complete or functional dentition and dental visits, frequency of tooth brushing and socio-demographic factors.

**Results:** In all age groups, the prevalence of dental visits and frequent tooth brushing increased and the prevalence of missing teeth decreased between 2002 and 2012. In 2012, the prevalence of a complete dentition was 87% in men and 85.3% women aged <25-yrs and 8.2% or 15.6% in the ≥85-yrs old. Prevalence of dental visits varied between 45.2% in the ≥85-yrs in 2002 and 73.1% in the 55-65-yrs old in 2012 in women and between 26.0% in the ≥85-yrs in 2002 and 68.1% in the 55-65-yrs old in 2012 in men. Frequent tooth brushing was more often reported by women (87.7%/81.4%) than men (73.5%/65.5%) in 2012 and 2002. Subjects aged ≥65-yrs, who visited their dentist within the last year, were twice as likely to have a functional dentition compared to subjects not having visited their dentist in the last year (men: 2.10, 1.68-2.63; women: 2.16, 1.73-2.70) in 2012, in 2002 this association was even stronger. A complete dentition was also associated with high income, higher education and non-smoking in 2012 in men and women.

**Conclusion:** In women and men, oral hygiene practices improved and the mean number of missing teeth substantially decreased between 2002 and 2012. Although women followed oral hygiene recommendations more closely than men, they still do not have a higher prevalence of a complete dentition, except in the oldest age groups.

## BACKGROUND

Gender differences in oral health have been documented across different cultures and across time.(1) Women usually report a higher number of missing teeth than men of the same age despite more intense oral hygiene and more frequent dental visits.(2) The association between oral health and gender is attributed to a multifactorial pathway through oral hygiene behaviours, social determinants, health care access and resources, diet and beauty ideals as well as through hormonal and reproductive factors.(3-5)

The number of missing teeth has been decreasing in past decades in many industrialized countries and the awareness of oral hygiene measures such as frequent tooth brushing has increased.(6, 7) Women, in particular, adopted oral hygiene recommendations and preventive check-up visits.(7-10) Within countries, there remain, however, differences between different sociodemographic groups: a higher number of missing teeth has been documented for those with lower education, low income, smokers or those who are obese.(11-14) These factors have also been associated with gender in many countries although differences changed during the past decades.(15-18) Thus, alterations in these factors may result in a changing pattern of missing teeth.

In women tooth loss increases with increasing parity, an observation also expressed in the proverb “a child, a tooth” which exists in different countries such as Germany, Russia and Japan.(19, 20) Pregnancy is associated with an increased risk of caries, and periodontal diseases, mostly related to increased predisposition to infection from higher progesterone levels, and changes in saliva composition with reduced pH-levels and buffer capacity.(21-23) In addition, women tend to postpone dental visits during pregnancy because of concerns about the safety of the unborn child, although current guidelines recommend dental visits prior to and during pregnancy.(24, 25) Promoting dental visits and monitoring of oral health during pregnancy and the decreasing number of children per women in industrialized countries such as Switzerland should thus be expected to contribute to eliminating gender differences in dentition.(25, 26)

In a nationwide survey assessing dental health in Switzerland in 2002, complete dentition was not associated with sex, although women were more likely to visit the dentist and to brush their teeth more frequently than men.(2) The aim of the current study was to assess the frequency of personal oral hygiene measures (visits to the dentist and tooth brushing) in women and men living in Switzerland in 2012, and to analyse whether these factors, as well as education, income, BMI and smoking, were associated with the dental status, and whether the associations with gender have changed since 2002.

## METHODS

### Data and study population

Data from the nationwide health surveys in 2002 and 2012 were analysed. In 2012, a representative sample of 41,008 private households was approached for participation. 21,597 participants took part in an initial telephone interview and 18,357 subjects filled in an additional written questionnaire. In 2002, the corresponding numbers were 30,824 private households, 19,706 telephone interviews, and 16,141 written questionnaires. Participants had to be at least 15 years old at the time of the interview. The questionnaire was provided in one of three native languages (German, French, or Italian). The analysis was restricted to the 17’784 subjects in 2012 and 14’661 subjects in 2002 who answered questions on missing teeth and restorations.

### Outcome

Descriptive analyses were conducted to quantify the mean number and prevalence of 1-2, 3-8, 9-27, or 28 (edentulous) missing teeth and dental restorations as well as the prevalence of oral hygiene practices. The question used to define the prevalence of missing teeth was: “How many teeth, excluding wisdom teeth, are missing from your mouth today? We have a maximum of 28 teeth without wisdom teeth.” If subjects answered having 0 missing teeth they were categorized as having a complete dentition; if they were missing up to 8 teeth they were categorized as having a functional dentition. The participants were asked whether they were wearing a prosthetic dental restoration; in detail they should indicate whether they were wearing crowns, one-piece post-and-core crowns, fixed dental prostheses (FDP, “bridges”), removable partial dentures, complete dentures, and/or dental implants. To reduce complexity, subjects were categorized into the following three groups: removable dental restoration (everybody indicating a removable partial denture or complete denture, irrespective of the presence of any fixed restoration or implants), fixed dental restoration (remaining subjects who indicated having crowns, one-piece post-and-core crowns, and/ or bridges), and implants-only (subjects indicating implants without specifying the dental restoration). Oral hygiene practices were assessed, firstly, by the question on how often subjects indicated visits to a dentist or dental hygienist in the last 12 months (“dental visits”) and those with positive answers to the question were grouped as visitors. Secondly, subjects were categorized into frequent tooth brushers, if they indicated to brush their teeth at least twice a day, otherwise they fell into the non-frequent group.

The outcomes of the logistic regression analyses were complete and functional dentition as defined above.

### Exposure

Descriptive analyses were conducted stratified by sex and 10-yrs age strata. Based on literature and on a previous analysis with data from 2002, exposures analysed in the logistic regression included dental hygiene practices (visits to the dentist in the last 12 months, frequency of tooth brushing) and socio-demographic factors.(2) Socio-economic status (SES) was assessed by asking subjects about their highest achieved level of education and household income. Education was then grouped into four categories: university degree (tertiary education), secondary education I (training on the job), secondary education II (general education), and compulsory or no/unknown education. Income quartiles were allocated as defined in the respective surveys (2002 and 2012): low (<2,750 Swiss francs (CHF) and <2,857 CHF]; low–middle (2,750–3,699 CHF and 2,857–3,999CHF); middle (3,700–4,999 CHF and 4,000–5,332 CHF); and high (≥5,000 CHF and ≥5,333 CHF). The differentiation into rural or urban was based on a population cut-off of at least 10,000 people or a group of communities close together with at least 20,000 people. Nationality was categorised into Swiss or Non-Swiss nationality. Self-reported height and weight were used to calculate the body mass index (BMI). BMI (kg/m^2^) was categorized into 5 groups: <18.5, 18.5-24.9, 25.0-29.9, >30.0, unknown. Smoking status was grouped into the following 4 categories: non-smoker, ex-smoker, current smoker and unknown.

### Statistical Analysis

The mean number of missing teeth and the prevalence of 1-2, 3-8, 9-27, or 28 missing teeth and the distribution of dental restoration (no restoration, removable restoration, fixed restoration, implants-only) were calculated, stratifying by sex and age. In addition, prevalence of “visits to the dentist and/or dental hygienist in the last year” and “frequent tooth brushing” were also stratified by sex and age. Prevalence rates were weighted with regard to age, gender, residential area and nationality to provide representative results for the Swiss non-institutionalized population.(26) The weights used accounted for sampling probabilities and missing information. Sex-stratified multivariate logistic regression analyses were performed to investigate associations between complete or functional dentition and visits to the dentist, tooth brushing and sociodemographic factors. The regression analyses were calculated separately for 2002 and 2012 and restricted to those ≥65 years, because the differentiation between complete and functional dentition is usually not relevant in younger age groups. The statistical analyses were conducted using SAS 9.4 and the level of significance was set at α = 0.05.

## RESULTS

The prevalence of oral hygiene practices and dental restorations stratified by sex and age are summarized in Table 1. In 2002 and 2012, women reported more often to have visited a dentist or a dental hygienist in the last year than men, and this difference was present across all age groups. However, in 2012 a considerably higher percentage of men and women above age 75 years, and even more above age 85, reported dental visits in the last year. Frequent tooth brushing was also more often reported by women (87.7%) than men (73.5%) in 2012, with highest differences in the age group of 74-84-yrs. Compared to 2002, the proportion of tooth brushing increased in all age groups except men above age 85, somewhat more pronounced in men in the middle age groups and in women aged 75-86-yrs.

**Table 1:** Prevalence of oral hygiene practices (dentist visits*, frequent tooth brushing) and dental restoration, stratified by sex and age, in 2002 and 2012

|  | Age<br>(years) |  |  |  |  |  |  |  |  |  |  |  |  |  |  |  | All |  |
| --- | --- | --- | --- | --- | --- | --- | --- | --- | --- | --- | --- | --- | --- | --- | --- | --- | --- | --- |
|  | <25 |  | 25-34 |  | 35-44 |  | 45-54 |  | 55-64 |  | 65-74 |  | 75-84 |  | 85+ |  |  |  |
| Prevalence of | 2002 | 2012 | 2002 | 2012 | 2002 | 2012 | 2002 | 2012 | 2002 | 2012 | 2002 | 2012 | 2002 | 2012 | 2002 | 2012 | 2002 | 2012 |
| **Visits to the dentist in the last year** |  |  |  |  |  |  |  |  |  |  |  |  |  |  |  |  |  |  |
| Men | 65.1 | 64.8 | 58.3 | 58.3 | 60.1 | 62.6 | 64.1 | 67.1 | 67.7 | 68.1 | 61.8 | 67.9 | 53.8 | 64.4 | 26.0 | 52.4 | 61.8 | 64.5 |
| Women | 71.7 | 68.6 | 63.8 | 60.7 | 68.5 | 66.6 | 71.0 | 71.4 | 70.9 | 73.1 | 64.6 | 70.8 | 48.8 | 66.3 | 45.2 | 56.6 | 66.7 | 68.1 |
| **Frequent\*\* Tooth Brushing** |  |  |  |  |  |  |  |  |  |  |  |  |  |  |  |  |  |  |
| Men | 68.1 | 72.6 | 69.3 | 74.3 | 65.2 | 77.9 | 65.1 | 74.6 | 63.2 | 70.9 | 61.8 | 73.2 | 59.7 | 64.8 | 72.4 | 70.5 | 65.5 | 75.5 |
| Women | 85.3 | 87.4 | 80.6 | 86.4 | 81.5 | 87.9 | 83.6 | 89.9 | 81.3 | 88.8 | 80.4 | 87.6 | 75.0 | 85.5 | 68.9 | 76.3 | 81.4 | 87.7 |
| **No PDR** |  |  |  |  |  |  |  |  |  |  |  |  |  |  |  |  |  |  |
| Men | 91.4 | 90.5 | 75.3 | 74.3 | 60.0 | 60.7 | 36.9 | 46.8 | 22.5 | 27.5 | 13.4 | 15.1 | 10.7 | 10.5 | 3.3 | 5.1 | 50.1 | 50.9 |
| Women | 90.7 | 93.5 | 78.9 | 83.0 | 58.7 | 68.5 | 30.7 | 48.9 | 14.8 | 22.7 | 10.8 | 10.6 | 4.6 | 7.3 | 2.9 | 2.7 | 46.0 | 51.2 |
| **Removable PDR** |  |  |  |  |  |  |  |  |  |  |  |  |  |  |  |  |  |  |
| Men | 0.4 | 1.2 | 1.5 | 1.7 | 3.8 | 2.8 | 16.7 | 5.5 | 27.9 | 17.0 | 47.0 | 27.8 | 68.0 | 43.6 | 80.0 | 61.2 | 16.4 | 10.8 |
| Women | 1.2 | 1.1 | 1.6 | 1.2 | 3.3 | 1.6 | 12.0 | 4.1 | 29.1 | 13.7 | 50.2 | 30.6 | 74.4 | 42.9 | 87.4 | 58.8 | 19.1 | 11.3 |
| **Fixed PDR** |  |  |  |  |  |  |  |  |  |  |  |  |  |  |  |  |  |  |
| Men | 7.8 | 6.9 | 22.0 | 19.0 | 35.0 | 31.9 | 45.4 | 44.4 | 49.1 | 52.2 | 39.4 | 54.2 | 21.1 | 42.6 | 16.8 | 33.8 | 32.1 | 34.8 |
| Women | 7.7 | 4.6 | 18.0 | 13.1 | 36.4 | 27.5 | 56.5 | 42.5 | 54.7 | 58.9 | 37.8 | 54.3 | 20.6 | 47.1 | 9.7 | 37.6 | 33.9 | 34.3 |
| **Implants-only** |  |  |  |  |  |  |  |  |  |  |  |  |  |  |  |  |  |  |
| Men | 0.3 | 1.4 | 1.2 | 5.0 | 1.1 | 4.6 | 0.9 | 3.2 | 0.5 | 3.3 | 0.2 | 2.9 | 0.2 | 3.4 | <0.1 | <0.1 | 0.8 | 3.5 |
| Women | 0.4 | 0.8 | 1.6 | 2.7 | 1.5 | 2.4 | 0.8 | 4.5 | 1.3 | 4.8 | 1.2 | 4.4 | 0.3 | 2.8 | <0.1 | 1.0 | 1.1 | 3.2 |
| **Implants-all** |  |  |  |  |  |  |  |  |  |  |  |  |  |  |  |  |  |  |
| Men | 0.3 | 1.5 | 1.3 | 6.3 | 2.6 | 7.0 | 3.4 | 8.8 | 4.9 | 12.9 | 5.6 | 18.1 | 4.1 | 20.0 | <0.1 | 16.5 | 2.9 | 9.4 |
| Women | 0.5 | 0.8 | 2.0 | 3.9 | 3.0 | 5.5 | 3.8 | 10.3 | 6.5 | 15.4 | 6.6 | 20.3 | 3.6 | 23.7 | 1.6 | 16.0 | 3.5 | 10.3 |

**Abbreviations:** PDR: any type of fixed or removable prosthetic dental restoration; implants only: without indication of fixed or removable restorations; implants all: sum of implants with indications of fixed or removable restorations or no such indication \*visits to a dentist or dental hygienist in the last 12 months; \*\* at least twice a day

In 2012, up to the age of 55-64-yrs, women had a lower prevalence of dental restoration than men, while in the older age groups they reported a higher prevalence of dental restorations. This pattern was driven by the prevalence of fixed restorations and implants-only. The prevalence of removable restorations was higher in men across all age groups except for the 65-74-yrs old. Compared to 2002 more people reported not having a dental restoration. This observation was more pronounced in women than in men and most pronounced in the middle-aged, which coincides with the lower number of missing teeth in 2012 compared to 2002 in the age groups 45-54-yrs and older (Fig. 1). There was an increase from 2002 to 2012 in the prevalence of implants across all age groups in men and women, the prevalence ranged between 0.8% in women <25-yrs and 23.7% in women 75-84-yrs. Implants were reported slightly more frequently by younger men than younger women, and somewhat more frequently by women aged 45-84 years than by men of the same age. The increase over time was most pronounced in men (4.1% to 20.0%) and women (3.6% to 23.7%) aged 75 to 84 years. The prevalence of implants in 2012 was highest in the highest income groups in men (12.5%) and in women (12.1%) and was slightly higher in women than in men across the other income groups (9.9%, 8.7%, 9.0% in women and 8.1%, 7.5%, 8.8% in men for income quartiles 1, 2 and 3, respectively).

**Figure 1.**
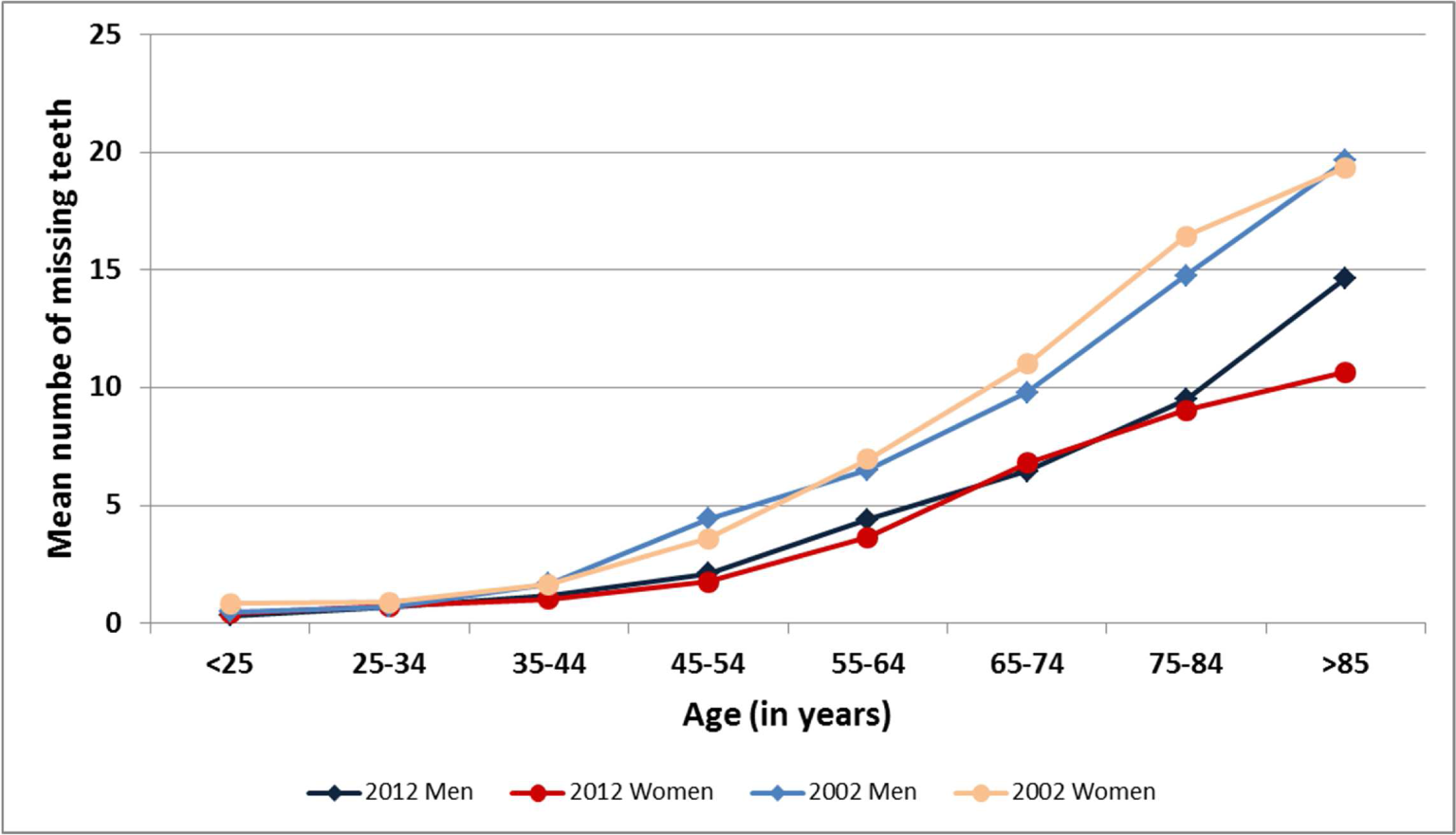
Mean number of missing teeth in 2002 and 2012, stratified by sex and age.

In 2012, the mean number of missing teeth in men and women was very similar up to the age group of 75-84-yrs, while in the oldest age group, the mean number of missing teeth was higher in men than in women (mean of 15 versus 11) (Fig. 1). In 2002, the mean number of missing teeth had been higher than in 2012 in all age groups except in the youngest, and in the 65-74 and 75-84-yrs old. A slightly higher number of missing teeth was reported by women than by men overall in both surveys (mean of 11 versus 10, and 16 versus 15, respectively). In those above age 85, it had been identical in 2002.

Figure 2 displays the prevalence of missing teeth in 2012 by sex and age. The overall prevalence of a complete dentition was slightly higher in men (52.7%) than in women (50.9%), decreasing strongly with age from 87.0% in men and 85.3% in women below 25-yrs to 8.2% of men and 15.6% of women aged ≥85-yrs. In 2002, the corresponding prevalence rates were 85.8% and 77.1% in subjects below 25-yrs, and 1.3% and 1.9% in male and female subjects ≥85-yrs, respectively (data not shown). On the other hand, the percentage of men and women with missing teeth was increasing strongly with age (Fig. 2). Compared to 2002, there has been a decrease in the prevalence in missing teeth across all age groups. In the whole population, women were more frequently missing 1-2 or 3-8 teeth (21.5%, 18.7%) than men (20.9%, 17.6%) in 2012, while the prevalence of a non-functional dentition (6.6% in men vs. 6.7% in women) or of edentulism (2.2% in both) was practically identical. Sex differences were most pronounced in the oldest age group, with 28.3% of men versus 34.5% of women reporting 9-27 missing teeth, and 22.7% of men versus 14% of women reporting edentulism.

**Figure 2.**
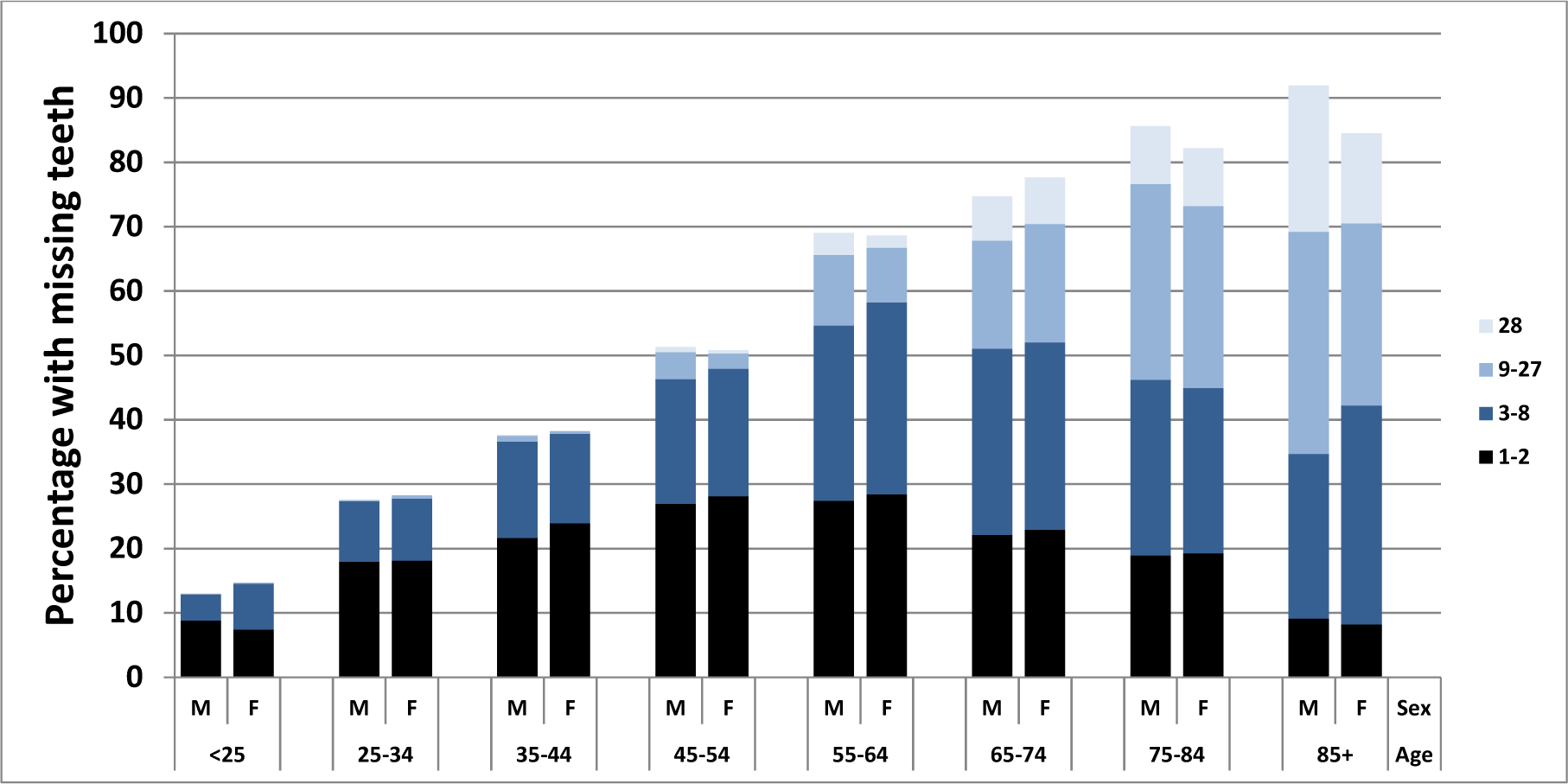
Prevalence of 1-2, 3-8, 9-27, or 28 missing teeth in 2012, stratified by sex and age. **Abbreviations:** M: men, F: women, Age in years

According to the logistic regression analyses, visits to the dentist over the last 12 months were significantly associated with complete dentition in women above age 65-yrs (OR 1.62, 95% CI 1.08-2.44) in 2002, while this association was no longer present in 2012. In men, this association was not observed, neither in 2002 nor in 2012 (Table 2). Subjects aged ≥65-yrs, who visited their dentist within the last 12 months, were however still twice as likely to have a functional dentition compared to subjects not having visited their dentist (men OR 2.10, 95% CI 1.68-2.63; women OR 2.16, 95% CI 1.73-2.7) (Table 3). These associations were less pronounced in 2012 than in 2002, both in men and women. In 2012, frequent tooth brushing (≥ 2x/d) compared to tooth brushing less than twice a day was neither associated with a complete dentition nor a functional dentition in subjects aged ≥65-yrs In women, it had even been associated with a decreased chance of having a complete dentition in 2002 (OR 0.63, 95% CI 0.41-0.96).

**Table 2:**
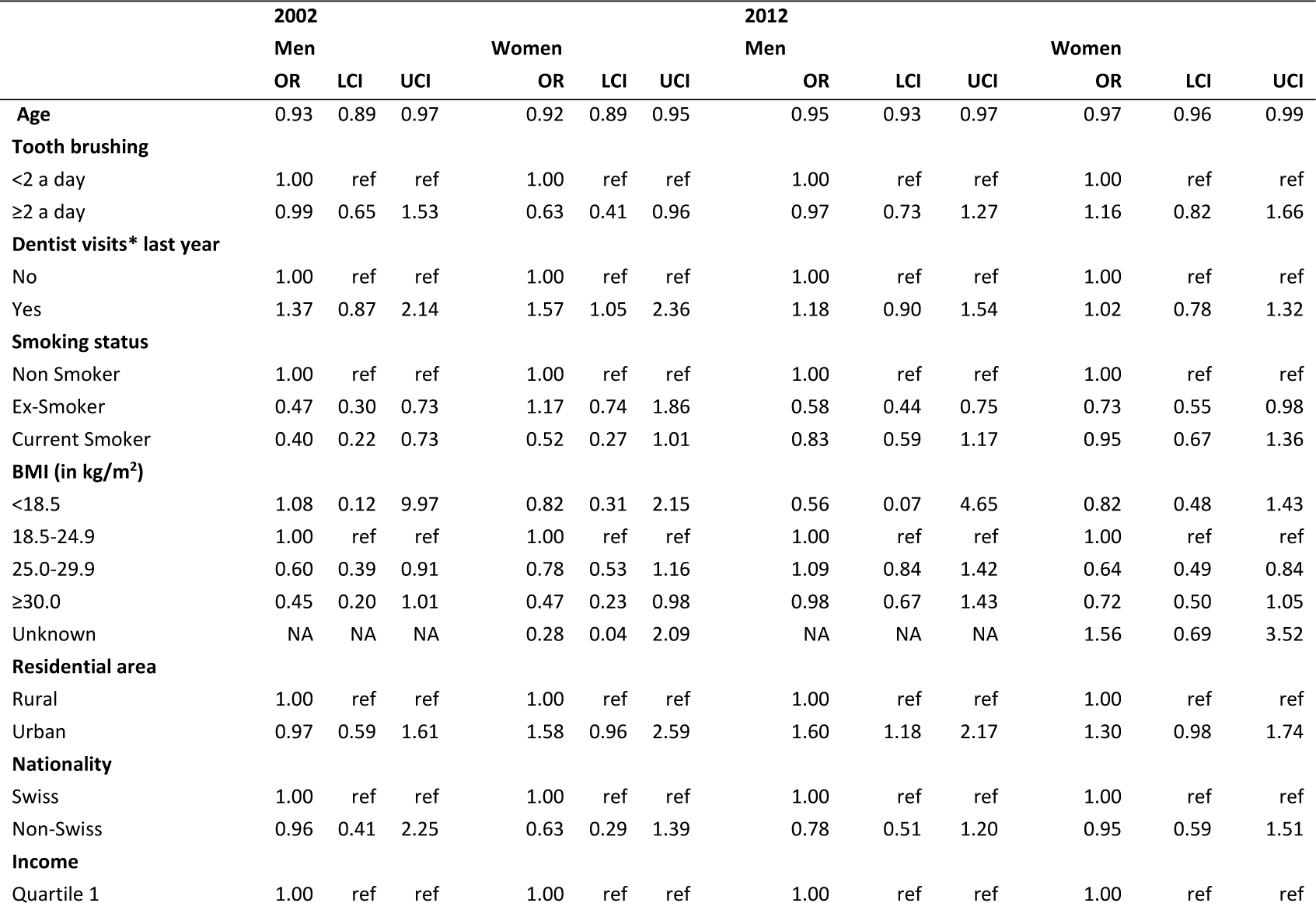
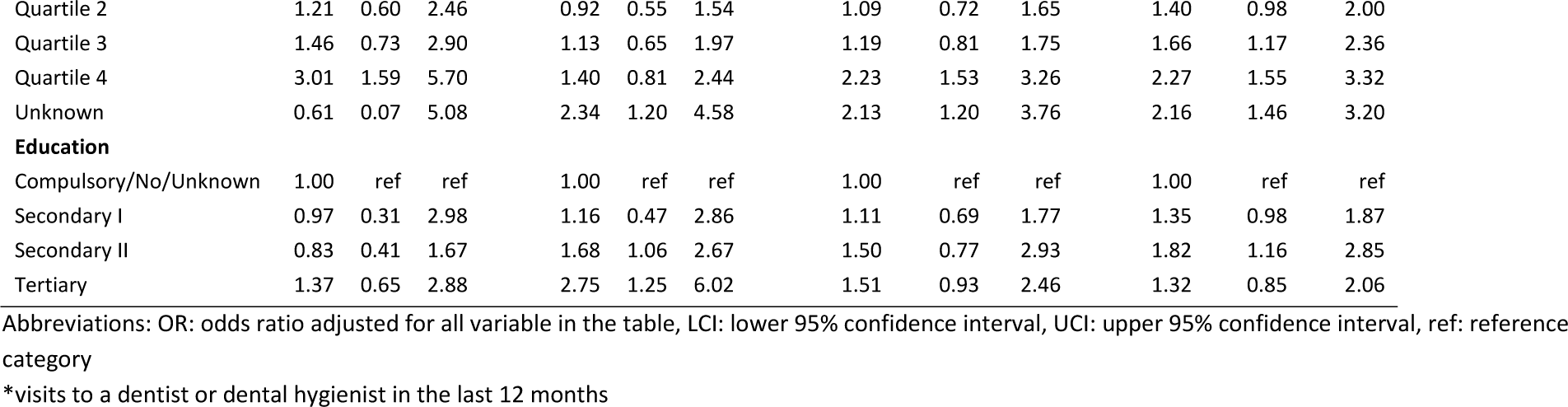
Association between complete dentition and sociodemographic factors in subjects ≥ 65 years, stratified by sex, in 2002 and 2012

**Table 3:** Association between functional dentition (up to 8 missing teeth) and sociodemographic factors in subjects ≥ 65 years, stratified by sex, in 2002 and 2012

|  | 2002 |  |  |  |  |  | 2012 |  |  |  |  |  |
| --- | --- | --- | --- | --- | --- | --- | --- | --- | --- | --- | --- | --- |
|  | Men |  |  | Women |  |  | Men |  |  | Women |  |  |
|  | OR | LCI | UCI | OR | LCI | UCI | OR | LCI | UCI | OR | LCI | UCI |
| **Age (continuous)** | 0.92 | 0.90 | 0.94 | 0.91 | 0.90 | 0.93 | 0.92 | 0.91 | 0.94 | 0.94 | 0.92 | 0.96 |
| **Tooth brushing** |  |  |  |  |  |  |  |  |  |  |  |  |
| <2 a day | 1.00 | ref | ref | 1.00 | ref | ref | 1.00 | ref | ref | 1.00 | ref | ref |
| $\geq 2$ a day | 1.03 | 0.79 | 1.34 | 0.94 | 0.72 | 1.23 | 0.97 | 0.76 | 1.23 | 0.85 | 0.62 | 1.15 |
| **Visits to the dentist last year** |  |  |  |  |  |  |  |  |  |  |  |  |
| No | 1.00 | ref | ref | 1.00 | ref | ref | 1.00 | ref | ref | 1.00 | ref | ref |
| Yes | 3.56 | 2.74 | 4.62 | 3.06 | 2.45 | 3.83 | 2.10 | 1.68 | 2.63 | 2.16 | 1.73 | 2.70 |
| **Smoking status** |  |  |  |  |  |  |  |  |  |  |  |  |
| Non Smoker | 1.00 | ref | ref | 1.00 | ref | ref | 1.00 | ref | ref | 1.00 | ref | ref |
| Ex-Smoker | 0.60 | 0.45 | 0.80 | 0.94 | 0.70 | 1.27 | 0.42 | 0.33 | 0.54 | 0.68 | 0.52 | 0.88 |
| Current Smoker | 0.35 | 0.24 | 0.50 | 0.60 | 0.42 | 0.85 | 0.40 | 0.29 | 0.57 | 0.51 | 0.37 | 0.71 |
| **BMI (in kg/m<sup>2</sup>)** |  |  |  |  |  |  |  |  |  |  |  |  |
| <18.5 | 0.75 | 0.16 | 3.51 | 1.58 | 0.91 | 2.75 | 0.33 | 0.08 | 1.36 | 0.79 | 0.48 | 1.31 |
| 18.5-24.9 | 1.00 | ref | ref | 1.00 | ref | ref | 1.00 | ref | ref | 1.00 | ref | ref |
| 25.0-29.9 | 0.75 | 0.57 | 0.99 | 0.98 | 0.77 | 1.25 | 0.96 | 0.75 | 1.23 | 0.73 | 0.58 | 0.93 |
| 30.0+ | 0.57 | 0.37 | 0.87 | 0.70 | 0.49 | 1.02 | 0.70 | 0.50 | 0.97 | 0.55 | 0.40 | 0.76 |
| Unknown | NA | NA | NA | 0.79 | 0.37 | 1.71 | 0.67 | 0.02 | 24.04 | 1.26 | 0.53 | 2.98 |
| **Residential area** |  |  |  |  |  |  |  |  |  |  |  |  |
| Rural | 1.00 | ref | ref | 1.00 | ref | ref | 1.00 | ref | ref | 1.00 | ref | ref |
| Urban | 0.92 | 0.68 | 1.25 | 1.62 | 1.24 | 2.11 | 1.45 | 1.13 | 1.85 | 1.40 | 1.09 | 1.78 |
| **Nationality** |  |  |  |  |  |  |  |  |  |  |  |  |
| Swiss | 1.00 | ref | ref | 1.00 | ref | ref | 1.00 | ref | ref | 1.00 | ref | ref |
| Non-Swiss | 1.16 | 0.66 | 2.02 | 1.41 | 0.91 | 2.18 | 1.24 | 0.86 | 1.78 | 1.15 | 0.76 | 1.72 |
| **Income (in CHF)** |  |  |  |  |  |  |  |  |  |  |  |  |
| Quartile 1 | 1.00 | ref | ref | 1.00 | ref | ref | 1.00 | ref | ref | 1.00 | ref | ref |
| Quartile 2 | 1.28 | 0.89 | 1.83 | 0.88 | 0.67 | 1.17 | 1.12 | 0.82 | 1.53 | 1.46 | 1.10 | 1.95 |
| Quartile 3 | 1.83 | 1.27 | 2.65 | 1.61 | 1.15 | 2.26 | 1.41 | 1.04 | 1.92 | 1.61 | 1.20 | 2.17 |
| Quartile 4 | 2.21 | 1.51 | 3.21 | 1.12 | 0.78 | 1.59 | 2.74 | 1.92 | 3.91 | 2.49 | 1.67 | 3.69 |
| Unknown | 0.31 | 0.11 | 0.94 | 1.75 | 1.15 | 2.67 | 1.53 | 0.92 | 2.54 | 1.56 | 1.09 | 2.22 |
| **Education** |  |  |  |  |  |  |  |  |  |  |  |  |
| Compulsory/No/Unknown | 1.00 | ref | ref | 1.00 | ref | ref | 1.00 | ref | ref | 1.00 | ref | ref |
| Secondary I | 1.64 | 0.78 | 3.44 | 4.51 | 2.60 | 7.83 | 1.43 | 1.02 | 2.00 | 1.56 | 1.21 | 2.01 |
| Secondary II | 1.16 | 0.78 | 1.71 | 2.36 | 1.83 | 3.03 | 2.25 | 1.22 | 4.18 | 2.37 | 1.50 | 3.75 |
| Tertiary | 1.67 | 1.06 | 2.63 | 2.94 | 1.68 | 5.14 | 2.01 | 1.37 | 2.94 | 2.79 | 1.77 | 4.38 |

Abbreviations: OR: odds ratio adjusted for all variable in the table, LCI: lower 95% confidence interval, UCI: upper 95% confidence interval

A complete dentition was furthermore associated with high income, higher education and non-smoking. The association between higher quartiles of income and complete dentition was most prominent in the elderly, with somewhat stronger associations for women in 2012 compared to 2002. A similar pattern appeared for higher education and complete dentition, while the associations for functional dentition were somewhat lower in 2012 for men. Current and ex-smoking men were less likely to have a complete dentition in 2002 and 2012, while in women, only ex-smoking was associated with a complete dentition. Current and ex-smoking was consistently associated with a lower likeliness to have a functional dentition for men and women in both surveys, with odds ratios for current smoking being below 0.5 for men.

In 2002, overweight and obese subjects were less likely of having a complete dentition compared to normal weight subjects. In 2012, this association was only observed for overweight women (OR 0.64, 95% CI 0.49-0.84) while in men chances of a complete dentition were similar in normal weight and overweight subjects (Table 2).

## DISCUSSION

Comparing oral hygiene measures with the number of missing teeth revealed that women do not have a better dentition than men although they brush their teeth more frequently. This is consistent with previous findings in Switzerland in 2002 (2), and also other countries such as Greece (7), Finland (8), Jordan (9), or Japan (10). Compared to earlier years the mean number of missing teeth has substantially decreased in women and men in Switzerland, particularly in the older population.(27) In 2012, the mean number of lost teeth was similar in men and women until the age of 64 years, while in the oldest age group, more teeth were missing in men than in women. This is in contrast to data from 2002, when women in the age groups 65-74 and 74-85-yrs had more missing teeth than men.

With regard to the prevalence of a complete dentition there remains a slight difference in favour of men for the total population; women were more frequently missing 1-8 teeth than men. This is again different in the age groups above 74 years, where the prevalence of a complete dentition is lower in men than in women, while the prevalence of edentulism is higher in men than in women. In the past, the prevalence of edentulism used to be higher in women than in men in many countries (28-31), but this has been changed in parts of the world over the past decades. With the decrease of the prevalence of edentulism, also the differences between men and women decreased.(28, 29, 32, 33)

Since 2002, oral hygiene practices have improved, possibly accounting for some of the improvements in dental status. Changes in dental care practices, possibly a more pronounced attention to the dental status and care of the elderly, may also have contributed to these improvements. We have observed an increase in prevalence of dental visits particularly in the older age groups while there was a small reduction in the younger age groups. The shift from fixed to removable restoration occurred about one age decade later in 2012 compared to 2002. A peak in fixed restorations of 52%-59% was found in the age groups 55–74-years in 2012, while this peak (45%-57%) was found in the group of 45-to 64-yrs-old subjects in 2002. A similar shift was observed for removable restorations, with a prevalence of 28-30% in the 65–74-yrs age group in 2012 and 28-29% in the 55–64-yrs age group in 2002. The increase in the prevalence of dental implants particularly in the age groups 45-yrs and older reflects the increasing indication for implants replacing missing teeth. The higher prevalence of implant restorations in higher income groups illustrates the association between financial limitations and therapeutic treatment options if payed out of the pocket. This association however only becomes apparent in middle aged and older men.

The current study revealed several associations between dentition and sex across sociodemographic factors. Elderly living in an urban setting were more likely to have a complete dentition than subjects living in a rural area. The association between higher quartiles of income and complete and functional dentition was somewhat stronger for women in 2012 compared to 2002. Lower education and income have repeatedly been associated with tooth loss in different countries.(11, 12, 27, 34) Smoking, which is often associated with a lower socio-economic status, was a strong risk factor for tooth loss, particularly in men: currently smoking men were less than half as likely to have a functional dentition than non-smokers. The association with BMI varied across years of the survey and sex, with overweight and obesity being negatively associated with a complete dentition in 2002 in men and women but only in women in 2012. Obesity has repeatedly been associated with periodontitis (35), which is a major reason for tooth loss in adults and associated with systemic inflammation. Obesity-associated inflammation and tooth loss might be differentially associated in men and women as indicated by studies focussing on different CRP levels and tooth loss.(13, 36) Obesity based on waist circumference was associated with tooth loss in pommerania, but in contrast to the current results the effects were stronger in men than in women.(36) Whether the observed association between higher body weight and tooth loss is mainly mediated by biological or social factors is not yet completely understood. The prevalence of obesity and overweight in Switzerland is higher in men than women and has slightly increased since 2002 in women and men. (37) Thus, the temporal trend is unlikely to account for the observed change in the gender difference. The same holds for smoking, and there is a slight decrease in the smoking prevalence overall from 2002 to 2012 in men and women, and a more pronounced decrease in heavy smoking (≥20 cigarettes per day) from 13.5% to 9% in men and from 7.3% to 4.3% in women (BFS 2016, page 10).

All results presented in the current study were based on self-reported information and the validity depends on correct report by the survey participants bearing a certain recall bias. Studies investigating the quality of self-reported number of missing teeth have reported a high accuracy of these self-reported information.(38, 39) As only subjects speaking one of the three native languages, who were contactable by telephone were eligible for this study, the results might not be completely representative for the Swiss population. To increase representativeness, the descriptive statistics were weighted taking into account sampling probabilities and missing information(26), nevertheless they may provide a somewhat too optimistic picture.

In conclusion, the current study demonstrated that the difference between men and women regarding oral hygiene and dental visits showed only minor decrease in 2012, and that the higher compliance to preventive oral health behaviour of women does not yet result in a higher prevalence of a complete dentition. Underlying mechanisms determining oral health in men and women might have a different impact on gender as the example of overweight indicated in this study and need further investigation.

## Data Availability

The data that support the findings of this study are available from the Swiss federal office of statistics but restrictions apply to the availability of these data, which were used under license for the current study, and so are not publicly available. Data are however available from the authors upon reasonable request and with permission of Swiss federal office of statistics.

